# Cost-effectiveness and health impact of screening and treatment of *Mycobacterium tuberculosis* infection among formerly incarcerated individuals in Brazil

**DOI:** 10.1101/2024.01.03.23300373

**Authors:** Ana van Lieshout Titan, Fayette Klaassen, Daniele Maria Pelissari, José Nildo de Barros Silva, Kleydson Alves, Layana Costa Alves, Mauro Sanchez, Patricia Bartholomay, Fernanda Dockhorn Costa Johansen, Julio Croda, Jason R. Andrews, Marcia C. Castro, Ted Cohen, Cornelis Vuik, Nicolas A. Menzies

**Affiliations:** Department of Global Health and Population, Harvard T.H. Chan School of Public Health, Boston, MA, USA; Delft Institute of Applied Mathematics, Delft University of Technology, Delft, The Netherlands; National Tuberculosis Programme, Ministry of Health, Brasilia, Brazil; Collective Health Institute, Federal University of Bahia, Salvador, Bahia, Brazil; Health and Environment Surveillance Secretariat, Ministry of Health, Brasilia, Brazil; Universidade Federal de Mato Grosso do Sul, Campo Grande, Brazil; Fiocruz Mato Grosso do Sul, Fundação Oswaldo Cruz, Campo Grande, Brazil; Department of Epidemiology of Microbial Diseases, Yale School of Public Health, New Haven, CT, USA; Division of Infectious Diseases and Geographic Medicine, Stanford University, Stanford, CA, USA; Center for Health Decision Science, Harvard TH Chan School of Public Health, Boston, MA, USA

## Abstract

**Background:** Formerly incarcerated individuals experience high tuberculosis (TB) incidence rates but are generally not considered among risk groups eligible for TB prevention. We investigated the potential health impact and cost-effectiveness of *Mycobacterium tuberculosis (Mtb)* infection screening and TB preventive treatment (TPT) for formerly incarcerated individuals in Brazil.

**Methods:** Using published evidence for Brazil, we constructed a Markov state transition model simulating TB-related health outcomes and costs among formerly incarcerated individuals. The analysis compared TB infection screening and TPT to no screening, considering a combination of *Mtb* infection tests and TPT regimens. We quantified health effects as reductions in TB cases, TB deaths and disability-adjusted life years (DALYs). We assessed costs from a TB programme perspective. We report intervention cost-effectiveness as the incremental costs per DALY averted, and tested how results changed across subgroups of the target population.

**Findings:** All TPT interventions were cost-effective in comparison to no screening, with a strategy including a tuberculin skin test and a 3-month isoniazid and rifapentine regimen costing $242 per DALY averted. It was estimated to avert 31 (95% uncertainty interval: 14-56) lifetime TB cases and 4.1 (1.4-8.5) lifetime TB deaths per 1,000 individuals receiving the intervention. Younger age, longer incarceration, and more recent prison release were each associated with significantly greater health benefits and more favorable cost-effectiveness ratios; however, the intervention was cost-effective for all subgroups examined.

**Interpretation:** *Mtb* infection screening and TPT appear cost-effective for formerly incarcerated individuals.

**Funding:** NIH.

**Evidence in context:** *Evidence before this study:* In many settings, incarcerated individuals have been shown to face higher risks of *Mycobacterium tuberculosis (Mtb)* infection than the general population. Individuals exiting prison have been found to experience elevated tuberculosis incidence rates over several years, and studies have also reported evidence of elevated tuberculosis incidence in surrounding communities. While several studies have investigated the health impact and cost-effectiveness of interventions to detect and prevent TB disease within prisons, few studies have examined the health impact and cost-effectiveness of interventions to treat *Mtb* infection among formerly incarcerated individuals.

*Added value of this study:* Using a Markov model, we simulated lifetime results among a cohort of formerly incarcerated individuals in Brazil offered screening and treatment for *Mtb* infection. To our knowledge, this is the first study to investigate the health impact and cost-effectiveness of screening and treatment among this cohort. The results contribute to the ongoing efforts to effectively reduce the TB burden and reach the WHO’s End TB goals in 2030.

*Implications of all the available evidence:* Screening and treatment of *Mtb* infection among formerly incarcerated individuals would produce substantial health benefits and be highly cost-effective in the setting examined in this study.

## Introduction

In many countries, prisons are high-risk locations for transmission of *Mycobacterium tuberculosis (Mtb)* and may pose additional risks for transmission to individuals in surrounding communities (1, 2). Prisons serve as amplifiers of tuberculosis (TB) transmission for several reasons. Living arrangements within prisons are often overcrowded and poorly ventilated, increasing the number of respiratory contacts between infectious and susceptible individuals (1, 3). Individuals entering prisons may have a higher prevalence of infectious TB disease than the general population, due to the shared socioeconomic determinants of TB and incarceration. Compared to the general community, incarcerated individuals may experience a greater concentration of risk factors for rapid TB progression, including undernutrition, untreated comorbid health conditions, smoking, and use of illicit drugs. Finally, in settings with limited prison health services, infectious individuals may experience greater delays before initiating TB treatment, extending the duration of infectiousness. As a result, high *Mtb* infection and TB disease incidence rates among incarcerated populations have been documented in many countries (4, 5).

Prison-based TB transmission is a particular challenge for countries in the Americas (6). In Brazil, the TB notification rate among incarcerated individuals is estimated to be 40 times greater than in the non-incarcerated population, with this difference widening between 2010-2019 (7). Combined with a growing prison population, total cases among incarcerated individuals increased by 40% between 2015-2022 (8, 9).

In addition to high incidence rates within prisons, prison-based TB transmission will also lead to many individuals exiting prison with recent *Mtb* infection. While data on TB incidence among formerly incarcerated individuals are not routinely reported, this number is likely to be substantial. The short duration of incarceration for the majority of prisoners, combined with the comparatively long incubation period for TB, means that many individuals will be at risk of progressing to TB disease after prison release (4, 10). These factors may contribute to elevated TB incidence among formerly incarcerated individuals. Within Brazil, elevated incidence rates among this group have been documented up to seven years after release (1), representing an ongoing health risk for these individuals as well as contributing to ongoing transmission in their communities (11).

The time between *Mtb* infection and the development of TB disease represents an opportunity for prevention. Brazil has adopted the WHO End TB Strategy, which provides a roadmap for accelerating global reductions in TB incidence and mortality, and has committed to eliminating TB by 2030 (12, 13). Expanding access to Tuberculosis Preventive Treatment (TPT) for populations at high risk of disease is a key component of the End TB strategy, and treating *Mtb* infection among formerly incarcerated individuals could reduce TB burden in this marginalized group as well as limit the impact of prison-based TB transmission on affected communities (14). Prior research has demonstrated the limited impact of mass screening for TB disease within prisons and prompted calls for the investigation of other potential control measures such as preventive screening and treatment (15).

In this study, we investigated the potential health impact and cost-effectiveness of TB prevention interventions among formerly incarcerated individuals. To do so, we synthesized the evidence on *Mtb* infection risks among incarcerated individuals in Brazil and used this to parameterize a mathematical model of TB outcomes following prison release. We used this model to simulate lifetime health outcomes under a range of intervention scenarios representing screening and treatment for *Mtb* infection, as compared to a base-case scenario representing no *Mtb* infection screening and passive TB diagnosis. We compared health benefits (quantified as disability-adjusted life years (DALYs)) to incremental costs to estimate intervention cost-effectiveness and examined the sensitivity of these results to changes in age at testing, duration of incarceration, and time since prison release.

## Methods

### Study model

We constructed a Markov state transition model simulating future health outcomes and intervention-related costs among a cohort of formerly incarcerated individuals (16). In this model, the study cohort is divided into compartments representing differences in TB health state and the receipt of treatment for *Mtb* infection and TB disease (Figure 1a). The model tracks individuals transitioning between these health states as a result of disease natural history and the initiation and discontinuation of treatment using a monthly timestep and records deaths from TB or non-TB causes. Upon initial infection, individuals enter one of several *Mtb* infection compartments designed to reproduce empirical trends in TB progression rates over time since infection (4, 17–21). Individuals progressing to TB disease can either be diagnosed and initiate TB treatment, self-cure (i.e., control the disease without treatment) or die from TB or background causes. Individuals treated for TB disease were assumed to receive a standardized 6-month treatment, following national guidelines (17). Individuals who recover from TB disease (by treatment or self-cure) face risks of reinfection, recurrent TB disease, or death due to background causes. We assumed that individuals with prior infection would have partial immunity against reinfection (20). Background mortality rates were based on general population life tables for Brazil (22).

**Figure 1:**
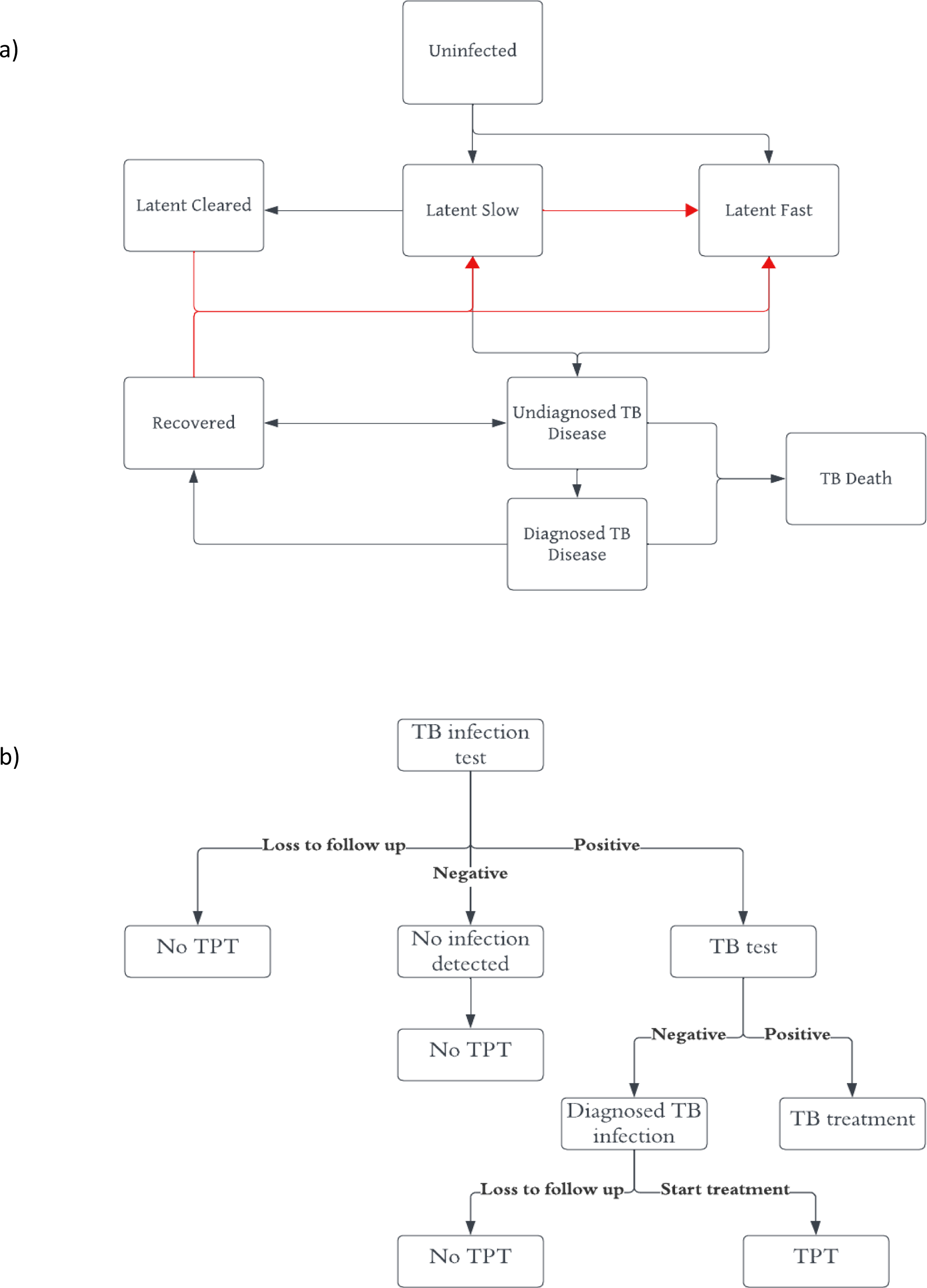
Schematic showing compartments and transitions of the Markov model (Panel A)* and decision tree for Mtb infection screening and treatment (Panel B). *Red arrows depict reinfection. All health states are subject to non-TB mortality. TPT = TB preventive treatment.

### Intervention scenarios

We constructed intervention scenarios representing a one-time screening for *Mtb* infection for individuals in the study cohort (Figure 1b). We modelled *Mtb* infection screening and treatment following WHO recommendations (23), with individuals who accept screening being tested for *Mtb* infection, and individuals testing positive subsequently screened for TB disease (24). We assumed that individuals diagnosed with *Mtb* infection but without TB disease would be offered TPT following a WHO-approved regimen. We included a probability of refusal of TPT. We assumed a fraction of individuals would discontinue TPT before completing the regimen, and included a probability of cure with partial treatment. We assumed that a fraction of those completing the regimen would clear *Mtb* infection. Cured individuals were assumed to face no further TB risk from their original infection, but could be reinfected. TPT was also assumed to confer protection against *Mtb* infection and progression to TB disease while individuals were receiving the regimen. Individuals diagnosed with TB disease were assumed to initiate TB disease treatment following national protocols (25). Individuals testing negative for *Mtb* infection were assumed to receive no further screening or treatment. We assumed the intervention would be offered to all individuals in the cohort except those being treated for TB disease. In addition to these intervention scenarios, we constructed a base-case scenario that assumed there would be no screening for *Mtb* infection. For all strategies, we assumed TB disease arising in the future would be identified and treated via passive diagnosis.

For the intervention scenarios we considered two different tests for *Mtb* infection (Tuberculin Skin Test (TST) and Interferon Gamma Release Assays (IGRA) (23)), and four TPT regimens (1 month of daily isoniazid and rifapentine (1HP), 3 months of weekly isoniazid and rifapentine (3HP), 4 months of daily rifampicin (4R), and 9 months of daily isoniazid (9H) (24)). We considered each combination of test and regimen.

### Model parametrization

Parameters describing TB epidemiology and natural history were estimated using a Bayesian evidence synthesis approach (26), with prior distributions for each parameter defined based on published values, and calibrated to reproduce available evidence on TB risks associated with incarceration. We validated the fitted model against published standards for TB natural history models (27). We used the RStan package (V2.21.8) (28, 29) to estimate epidemiological parameters. Fitted values for these parameters are provided in Table 1 and additional details in Tables S1 and S2. Parameters defining test sensitivity and specificity, regimen efficacy and discontinuation rates, and costs for each intervention component, were based on published studies (Table 1).

**Table 1:**
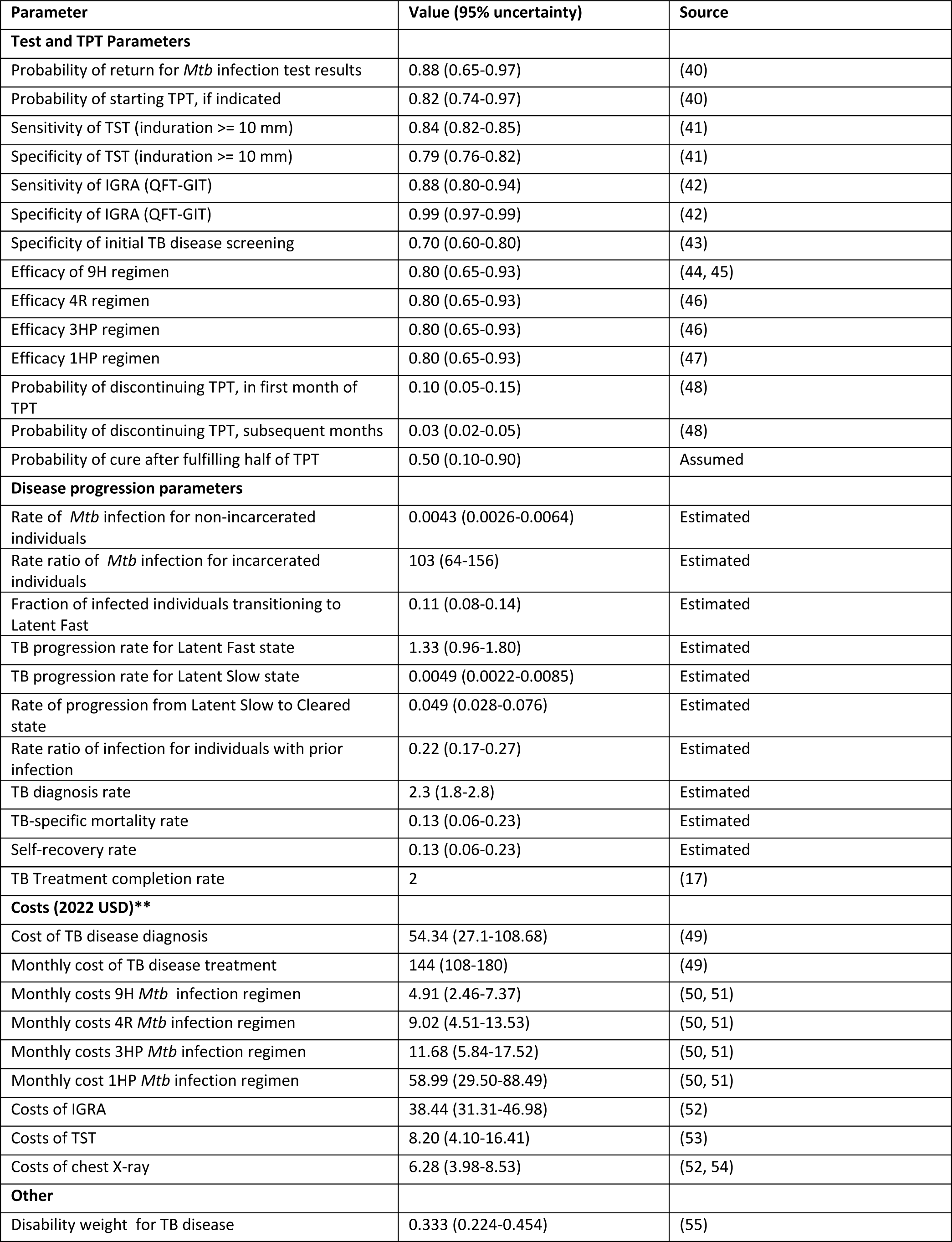
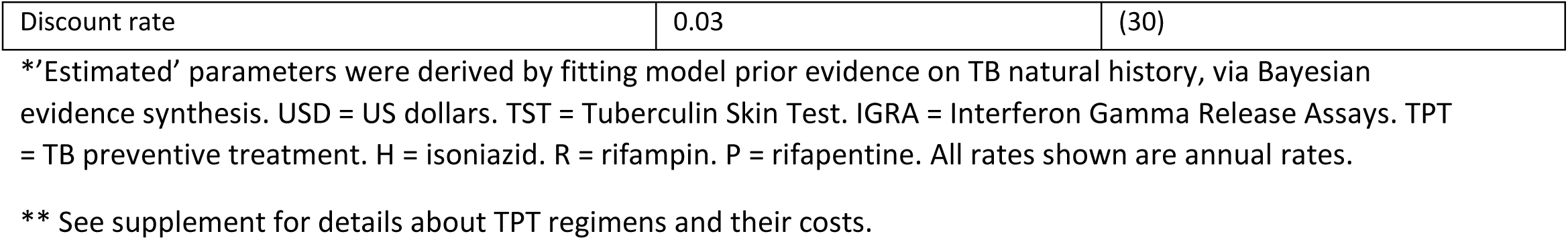
Parameter definitions, values and sources.

### Study outcomes

We quantified health benefits as reductions in TB-attributable DALYs, comparing each intervention scenario to the base-case. We also calculated the number of TB cases and TB deaths averted. We reported DALYs, TB cases, and TB deaths per 1,000 people. We estimated incremental costs in 2022 US dollars ($) from a TB program perspective, based on changes in diagnosis and treatment costs between intervention scenarios and the base-case. We estimated all outcomes over the lifetime of the study cohort.

We estimated incremental cost-effectiveness ratios (ICERs) to describe the cost-effectiveness of each intervention approach. Cost-effectiveness results are reported as the incremental cost per DALY averted, with costs and health outcomes discounted using a 3% discount rate (30). In sensitivity analyses, we re-estimated ICERs with the undiscounted costs and health outcomes (Figure S3). An intervention was deemed cost-effective in comparison to another intervention if the ICER fell under the local cost-effectiveness threshold. This was defined as 71-109% of current GDP per capita, based on estimates of the opportunity cost of healthcare spending (31). This implied a threshold falling in the range of $6,300-9,700 per DALY averted, based on a 2022 Brazilian GDP per capita of $8917 (32). Interventions were dominated if they had higher costs and worse health outcomes compared to other available strategies. Uncertainty in the ICERs was taken into account in the cost-effectiveness acceptability curves (CEACs). We conducted all analyses in R (V4.2.3) (33).

### Analysis of age, duration of incarceration, time since release

For the main analysis, we assumed that the cohort offered screening would be thirty years of age, had completed two years in prison, and were released from prison three months before the intervention was offered. We conducted additional analyses to understand how each of these factors (age at screening, duration of incarceration, and time since release) would affect intervention health impact and cost-effectiveness. To do so, we adjusted each factor over a range of plausible values (holding the other two factors at their original values) and recalculated study results.

We varied age at screening between ages 25 and 65, duration of incarceration between 1 month and 10 years and time since release from zero months to 10 years (Table S9).

### Statistical analysis

We propagated uncertainty through the analysis using second-order Monte Carlo simulation (34). To do so we specified probability distributions for each uncertain parameter (Table 1) and used these distributions to sample 1,000 values for each parameter. We recalculated study outcomes for each of these parameter sets, yielding a distribution of results for each outcome. We summarized these distributions as the mean and 95% uncertainty interval, representing the uncertainty in each study outcome due to the combined uncertainty in model parameters.

### Sensitivity analyses

We conducted one-way deterministic sensitivity analyses testing how changes in each parameter affected cost-effectiveness results. To do so, we recalculated results while varying each parameter between the ranges shown in Table 1, holding other parameters at their mean value. We conducted an additional sensitivity analysis to understand how elevated future reinfection rates (as could happen with reincarceration) change cost-effectiveness results. To do so, we specified an elevated value for the TB force of infection after prison release, operationalized as a rate ratio applied to the force of infection in the general population, and varied this rate ratio between 2 and 50. This rate ratio was assumed to decline linearly to 1 over the twenty years after release, to represent a declining probability of reincarceration with greater time since release (Figure S2). We also calculated CEACs (35) to report how the probability of each strategy being cost-effective changed across the range of the cost-effectiveness threshold.

## Results

### Health impact and cost-effectiveness of intervention strategies

As estimated by the Bayesian evidence synthesis, 2.4% (1.8-3.1) of the starting cohort was estimated to have TB disease and 57.1% (42.5-73.3) to have *Mtb* infection (Table S3).

Under the base-case scenario (no screening and TPT), we estimated there would be 120 (95% uncertainty interval: 75-181) TB disease cases, 13 (5–25) TB deaths, and 432 (186–772) DALYs per 1,000 persons (undiscounted) when estimated over the lifetime of individuals in the study cohort. Each of these outcomes was reduced under the interventions scenarios, with health impact ranging from 28 (13–52) TB cases, 3.7 (1.4-7.7) TB deaths, and 126 (49–246) TB DALYs averted per 1,000 under the TST/9H scenario, up to 34 (16–62) TB cases, 4.5 (1.5-9.4) TB deaths, and 149 (55–292) TB DALYs averted under the IGRA/1HP scenario (a 28% (18–38), 33% (22–43), and 30% (6–49) reduction in lifetime TB cases, deaths and DALYs respectively, as compared to the base-case). In general, greater health gains were produced under scenarios using IGRA instead of TST, and with shorter TPT regimens instead of longer regimens (Table 2).

**Table 2:**
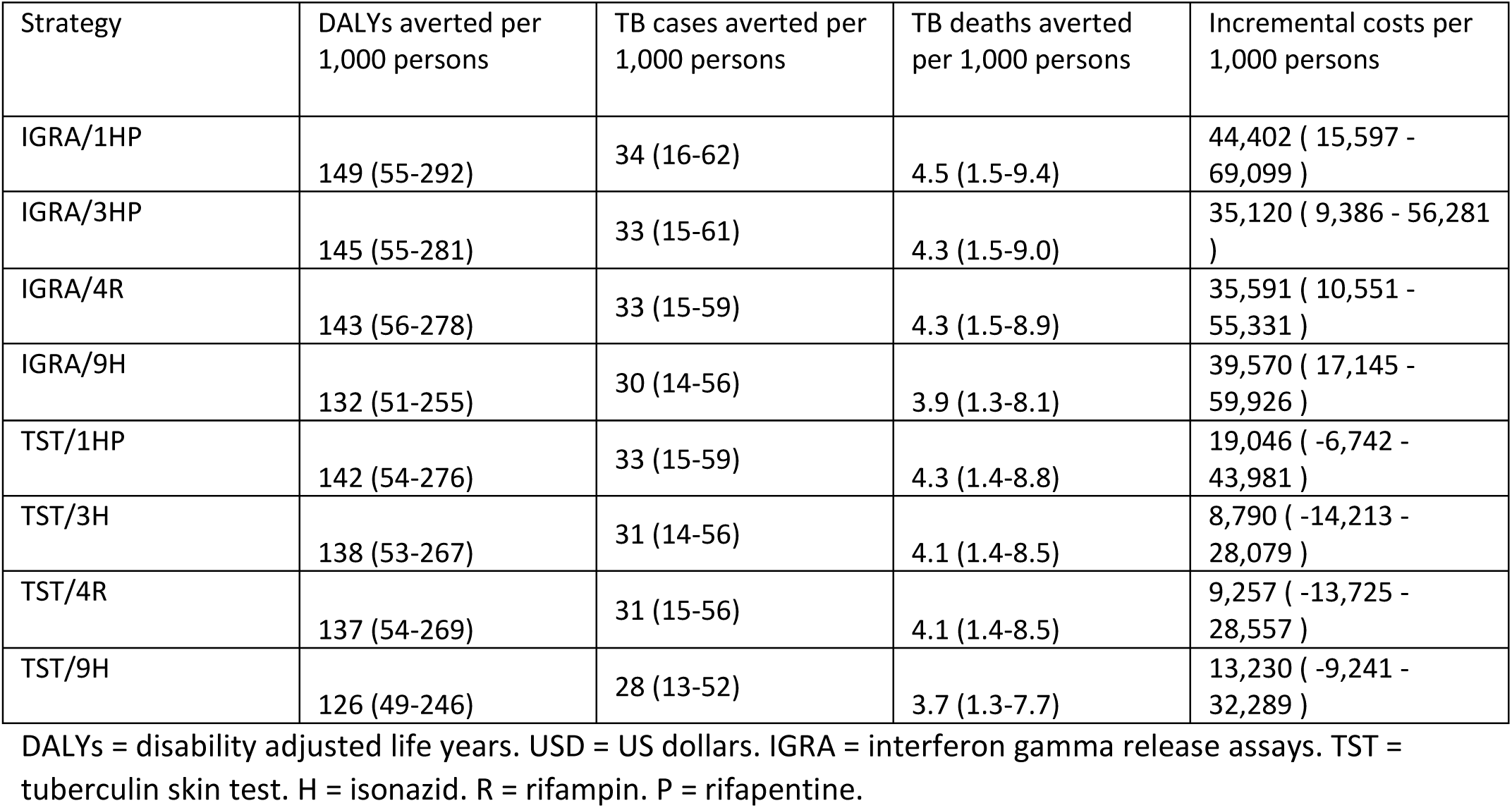
Incremental lifetime health benefits and costs for each intervention scenario, as compared to the base-case scenario.

Undiscounted lifetime costs were $99,585 (58,552–157,598) per 1,000 persons under the base-case scenario. Costs were higher under the intervention scenarios, with incremental costs (compared to the base case) ranging from $8,790 (ࢤ14,213–28,079) for the TST/3HP scenario up to $44,402 (15,597–69,099) for the IGRA/1HP scenario. Incremental costs were higher for scenarios employing IGRA instead of TST, and using 9H and 1HP as the TPT regimen versus 3HP or 4R (see Table 2).

Figure 2a shows the results of the cost-effectiveness analysis. Intervention strategies including, 9H or 4R, andIGRA/3HP were dominated by TST/1HP, TST/3HP and IGRA/1HP strategies. Compared to the base-case scenario, TST/3HP had an ICER of $242 per DALY averted. TST/1HP was both more effective and costly, with an ICER of $5,569 per DALY averted compared to TST/3HP. When TST/1HP was compared to the base-case scenario (for example, if the 3HP regimen were not available) the ICER was $383 per DALY averted. IGRA/1HP had an ICER of $7,066 compared to TST/1HP. Comparing these results to the cost-effectiveness threshold ($6,300-9,700 per DALY averted) showed that all strategies would be cost-effective compared to the base-case. For the optimal test-regimen combination, the choice between TST/1HP and IGRA/1HP was inconclusive, with TST/1HP preferred with a lower cost-effectiveness threshold (i.e., when the opportunity cost of spending is higher), and IGRA/1HP preferred with a higher threshold. See Supplement for more details.

The incidence rate five years after prison release was estimated to be 225 (153–485) per 100,000 under the base-case and 184 (97–308) per 100,000 with TST/3HP (Figure S1).

### Age, duration of incarceration, and time since release

We conducted additional analyses to understand how age at screening, duration of incarceration, and time since release affected intervention outcomes. For these analyses, we used TST/3HP as the intervention strategy. When we varied age at screening we found that health benefits (DALYs averted) were greatest and the ICER lowest for younger individuals. ICERs ranged from $216 per DALY averted for individuals aged 25 years at screening up to $1,081 for individuals aged 65 years. When we varied the duration of incarceration, we found that health benefits were greatest and ICERs lowest for cohorts with a longer duration of incarceration, although this trend plateaued for durations greater than 2 years. ICERs ranged from $243 per DALY averted for a 2-year duration of incarceration up to $1,716 for a 1-month duration of incarceration. Lastly, we varied the delay between prison release and the intervention being offered. Health benefits were greatest and ICERs lowest with testing soon after release, with both immediate screening and a 1-month delay having ICERs around $208 per DALY averted. ICERs increased with longer delays, with an ICER of $776 per DALY averted estimated for a cohort tested 10 years after prison release. Results for these analyses are shown in Figure 2b-d, and detailed results are shown in Table S9.

**Figure 2:**
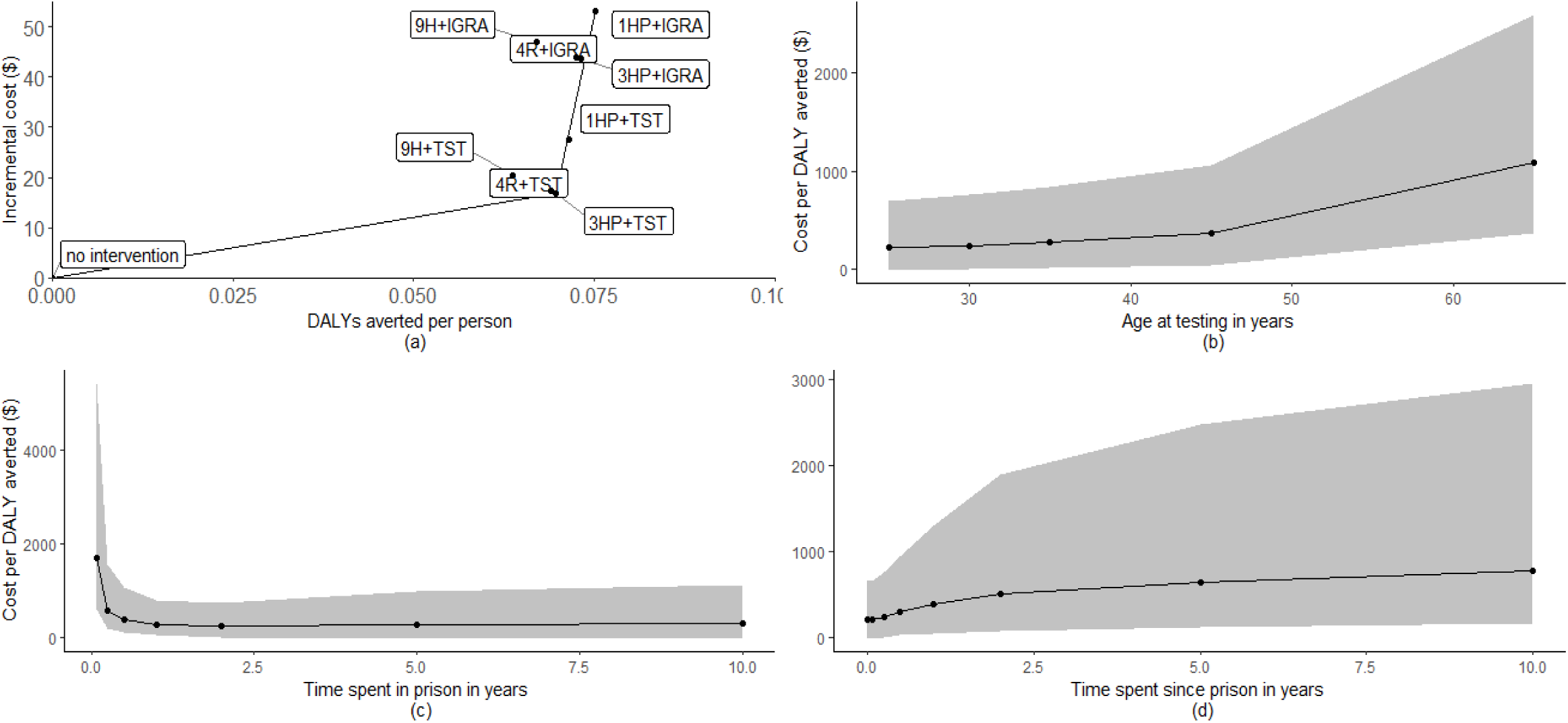
Cost-effectiveness plane of interventions and basecase scenarios (Panel A). Change in ICER for TST/3HP versus basecase produces by changes in age at testing (Panel B), duration of incarceration (Panel C), and time since prison release (Panel D). Shaded area represents 95% uncertainty. DALY = disability adjusted life-years.

### Sensitivity analysis

For a cost-effectiveness threshold below $300 per DALY averted, the base-case scenario of no screening and treatment had the highest probability of being cost-effective (Figure 3a). For a threshold between $300 and $7,100 per DALY averted, TST/3HP had the highest probability of being cost-effective and for a threshold above $7,100 per DALY averted IGRA/1HP had the highest probability of being cost-effective. There was substantial uncertainty about the best test-regimen.

**Figure 3:**
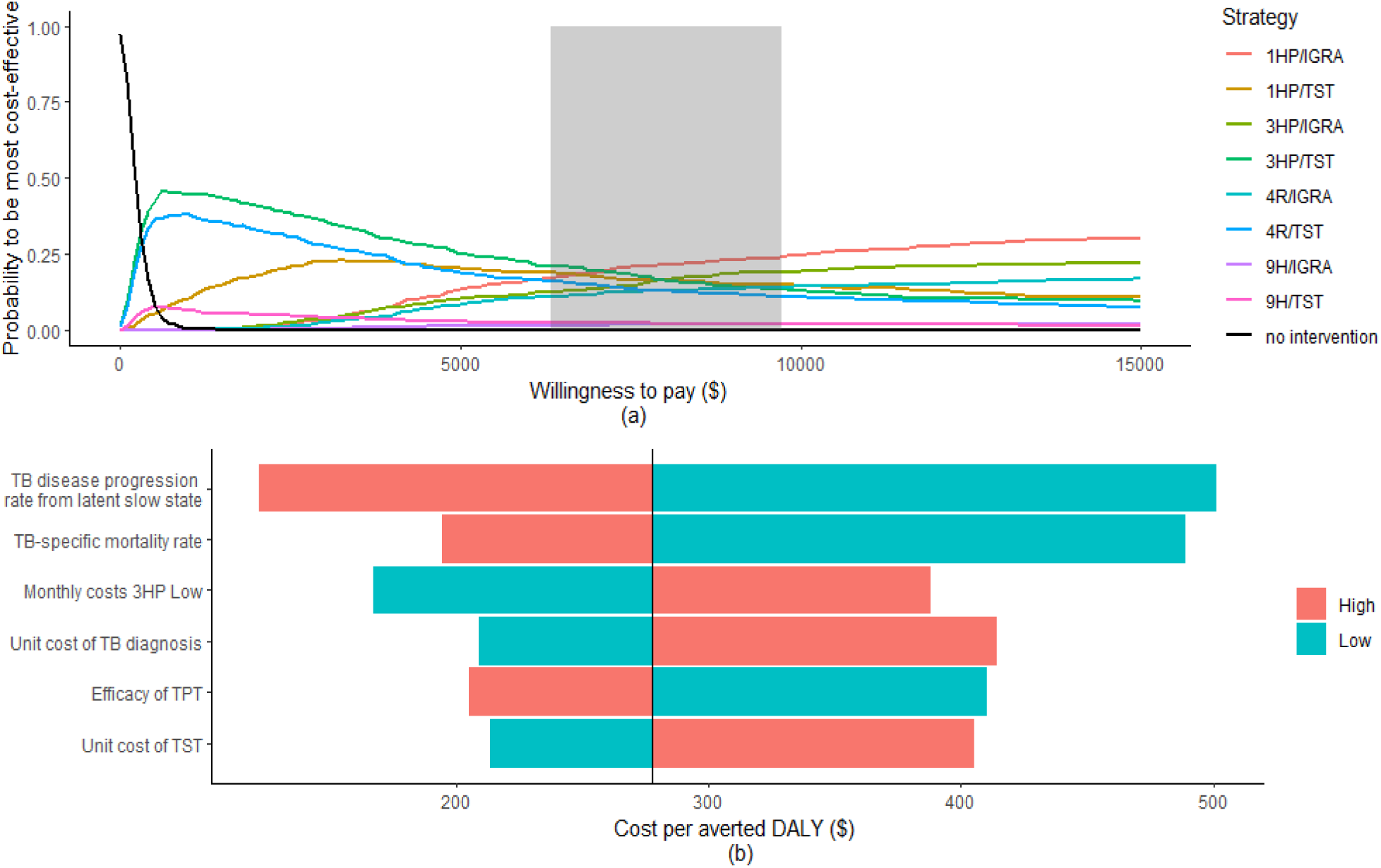
a) Cost-effectiveness acceptability curve for base-case and intervention scenarios. Grey-shaded area represents cost-effectiveness threshold in Brazil of 71-109% of the GDP per capita in 2022. b) Tornado diagram showing the six most influential parameters as calculated in the one-way deterministic sensitivity analysis. TST = tuberculin skin test. IGRA = interferon gamma release assays. TPT = TB preventive treatment. H = isonazid. R = rifampin. P = rifapentine. DALY = disability adjusted life years.

In one-way deterministic sensitivity analyses the parameters with the greatest influence on the ICER for TST/3HP versus the base-case were the rate of progression to TB from the Latent Slow compartment, TB-specific mortality, the monthly cost of the TPT regimen, the unit cost of TB diagnosis, the efficacy of the TPT regimen, and the cost of TST (Figure 3b), although none changed the conclusion of our analyses. We also tested how elevated reinfection rates after the intervention would affect the cost-effectiveness results. In this analysis elevated reinfection rates led to higher ICERs, but even with a reinfection rate 50 times higher than the general population the ICER for TST/3HP vs. the base-case was $303 per DALY averted, still well below the cost-effectiveness threshold.

## Discussion

In this study, we examined the potential health impact and cost-effectiveness of *Mtb* infection screening and TPT for formerly incarcerated individuals in Brazil. We found that the intervention would be highly cost-effective in this setting, with a cost per DALY averted estimated to be less than one-tenth of the cost-effectiveness threshold adopted for this analysis. This result was robust to uncertainty in input parameters and was true for all subgroups examined. Across all scenarios, TST/3HP, TST/1HP and IGRA/1HP were all highly cost-effective compared to the base-case scenario, but the analysis was inconclusive as to which strategy should be preferred at the suggested cost-effectiveness threshold.

Among the test and regimen options examined, we found that scenarios using IGRA for *Mtb* infection testing produced greater health benefits compared to those using TST, which is consistent with published research (36). The analysis also found that shorter TPT regimens were more effective than longer regimens. Shorter regimens have lower default rates, and were, in this study, assumed to have similar efficacy. Therefore, shorter regimens had greater health effects and were more cost-effective (37, 38).

We also analyzed how age at testing, duration of incarceration, and time since release affected study results. We found that the health impact of the intervention was greater for younger individuals, with a greater life expectancy over which TB could develop, and a greater number of life years lost when TB death occurs at a younger age.

Health benefits were also estimated to be greater for individuals with a longer duration of incarceration with these individuals more likely to acquire infection before release, and therefore having more to gain from the preventive intervention. This effect plateaued for durations of incarceration greater than two years.

Finally, we found that health benefits were greatest for individuals tested soon after prison release. As the risk of developing TB declines with increasing time since infection, individuals tested shortly after release are more likely to have recent infection and experience high risk of TB in the absence of TPT. For each of these subgroup analyses, greater health benefits were associated with lower (i.e., more favorable) cost-effectiveness ratios.

This study has several limitations. First, we did not explicitly simulate reincarceration among the target population. An average of 21% of formerly incarcerated individuals return to prison within the first year of release, increasing to 38.9% after five years since release (39). The reincarceration could lead to re-infection among individuals who had received TPT, reducing intervention benefits. We tested how study results would be affected by higher re-infection rates, and found that cost-effectiveness conclusions were robust to re-infection rates up to 50x higher than those assumed in the main analysis. Reincarceration could also reduce the fraction of individuals completing the TPT regimen. While higher rates of discontinuation were associated with lower health benefits, the intervention was found to be cost-effective across the range of discontinuation rates examined in this study. Second, there was substantial uncertainty around several parameters in the analysis. Because this uncertainty is included in the cost-effectiveness conclusions, the overall intervention effects were estimated imprecisely with wide uncertainty intervals. Third, it is possible that individuals accepting the intervention would differ systematically from the overall population of formerly incarcerated individuals. We did not investigate these selection effects. It is possible that these selection effects could promote cost-effectiveness (for example, if individuals choosing to be screened are more likely to accept and complete TPT), but they could also have the opposite effect (if individuals choosing to be screened are systematically healthier or with lower infectious exposure during prison). This represents an additional unmodelled source of uncertainty in the study results.

While this study found the intervention to be highly cost-effective, it is possible that uptake would be lower than assumed among the target population, given that *Mtb* infection does not produce symptoms that might motivate an individual to seek screening, and the benefits of avoiding future disease risks may not have great saliency for these individuals. Individuals might avoid any linkage to incarceration due to social stigma, impacting the acceptance of treatment. Given the highly favorable cost-effectiveness ratios, additional resources could be devoted to improving acceptance and completion rates without compromising intervention cost-effectiveness. We did not account for reduction in TB transmission, which could further benefit both the marginalized population and the surrounding communities. Considering the high recurrence rate, reduction in TB transmission among formerly incarcerated individuals might also affect the TB transmission risks in prisons.

In summary, TB prevention among formerly incarcerated individuals has received little attention, but this population is large and experiences a high burden of TB disease. The findings of this study suggest that screening and treatment of *Mtb* infection would be a cost-effective and impactful intervention for this population.

## Data Availability

All data produced in the present work are contained in the manuscript

