## Supplement Tables and Figures for "Cost-effectiveness and health impact of screening and treatment of *Mycobacterium tuberculosis* infection among formerly incarcerated individuals in Brazil"

### Table of contents

**Supplementary methods**......pg 2

**Methods for Bayesian parameter estimation**......pg 2

**Methods for DALY calculation**......pg 2

**Supplementary parameters and information**......pg 3

**Distribution of cohort across health states at the point of testing**......pg 3

**Validation check Markov model**......pg 3

**Supplementary exhibits**......pg 4

**Sensitivity Analysis**......pg 6

### Supplementary Methods

#### Methods for Bayesian parameter estimation

The Bayesian estimation model followed the same structure as the main study model and was used to simulate TB exposure before and during incarceration to represent the effect of incarceration on *Mtb* infection and progression. We ran this model from birth to age 35 and assumed entry to prison at age 20. We specified prior distributions for all model parameters (Table S1) and calibrated the model to reproduce a TB incidence rate in prison of 1600 per 100,000, a prevalence of *Mtb* infection prior to prison of 7.6% and in prison of 25% (1, 2) (Table S2). We implemented this analysis using the Stan package in R, running the model for 1250 iterations on 4 chains, keeping the last 250 samples of each chain. This provided us with 1,000 samples from the posterior distribution for each parameter, which we used in the main analysis. We report the mean estimate and 2.5% and the 97.5% quantiles for each parameter in Table 1.

**Table S1: Prior distributions for estimated epidemiological parameters**

| Parameter | Prior (95% uncertainty) | Distribution | Source |
|---|---|---|---|
| Rate of *Mtb*infection for non-incarcerated individuals | 0.10 (0.0041-0.29)* | Half normal | Assumed |
| Rate ratio of *Mtb* infection for incarcerated individuals | 31.9 (1.26-89.7)* | Half normal | (3) |
| Fraction of infected individuals transitioning to Latent Fast state | 0.03 (0.01-0.05) | Beta | (4-6) |
| TB progression rate for Latent Fast state | 1.00 (0.75-1.5) | Gamma | (4-6) |
| TB progression rate for Latent Slow state | 0.004 (0.002-0.007) | Gamma | (4-6) |
| Rate of progression from Latent Slow to Cleared state | 0.05 (0.03-0.08) | Gamma | (4-6) |
| TB diagnosis rate | 2.00 (1.50-2.50) | Gamma | Assumed |
| Rate ratio of infection for individuals with prior infection | 0.21 (0.15-0.25) | Beta | (7) |
| Self-recovery rate | 0.15 (0.05-0.25) | Gamma | (8) |
| TB-specific mortality rate | 0.15 (0.05-0.25) | Gamma | (8) |
| TB Treatment completion rate | 2.0 (1.6-2.3) | Gamma | Based on assumed 6 months disease duration |
| Prison exit rate | 0.5 (0.1-1) | Gamma | Assumed |
| Recurrence rate | 0.004 (0.002-0.007) | Gamma | Assumed |

*The half-normal distribution was implemented in stan as a normal distribution with mean 0 and standard deviation following the distribution.

**Table S2: Calibration targets and estimated value for the Bayesian model**

| Parameter | Value from literature | Estimated value with 95% posterior interval |
|---|---|---|
| TB disease incidence in prison (per 100,000) | 1600 (1500-1700) | 1401/100,000 person-years (1207-1603) |
| Prevalence *Mtb* infection prior to prison | 7.6% (3-14) | 10% (7-14) |
| Prevalence *Mtb* infection in prison | 25% (17-34) | 40% (33-48) |

#### Methods for DALY calculation

The following calculations were used to calculate Years Lived Disabled (YLDs), Years of Life Lost (YLLs), which are summed to calculate DALYs:

$$YLD = \frac{1}{12}\sum_{i}\sum_{m} N_{i,m} * w_i * (1+d)^{\frac{1-m}{12}}$$

Where $w_i$ is the disability weight associated with health state $i$, $N_{i,m}$ is the amount of life-months $m$ spent in health state $i$, and $d$ is the discount rate of 3%.

$$YLL(d > 0) = \sum_{x}\sum_{m} D_{x,m}\frac{(1-(1+d)^{-e_x})}{d} * (1+d)^{\frac{1-m}{12}}$$

$$YLL(d = 0) = \sum_{x}\sum_{m} D_{x,m} * e_x$$

Where $D_{x,m}$ is the amount of deaths in month $m$ at age $x$, $e_x$ is the life expectancy at age *x* according to Global Burden of Disease reference life table. We calculated YLL in a scenario with no TB ($YLL_{notb}$). Then we calculated the YLL due to TB:

$$YLL_{TB} = YLL - YLL_{notb}$$

And then we can calculate the DALY:

$$DALY = YLL_{TB} + YLD$$

**Supplementary parameters and information**

**Distribution of cohort across health states at the point of testing**

The initial values were calculated taking the distribution of the cohort at thirty years, after a duration of incarceration of two years, and released three months prior to being offered *Mtb* screening.

**Table S3: Distribution of the target population across modelled health states at point of screening**

| **Health state** | **Mean value (95% uncertainty)** |
|---|---|
| Uninfected | 0.367 (0.230-0.498) |
| Latent fast | 0.017 (0.010-0.025) |
| Latent slow | 0.486 (0.376-0.603) |
| Latent cleared | 0.068 (0.039-0.105) |
| Undiagnosed TB disease | 0.012 (0.009-0.016) |
| Diagnosed TB disease | 0.012 (0.009-0.015) |
| Recovered | 0.038 (0.028-0.049) |
| Dead (other causes) | 0 |
| Dead (TB cause) | 0 |

**Validation check Markov model**

The Markov model was validated to the values described in Table S4, following the natural progression (9).

**Table S4: Validation targets used for the Markov model and the estimated value of the target calculated with the model**

| **Parameter** | **Value from McQuaid** | **Estimated value in model** |
|---|---|---|
| The cumulative incidence over the first five years of infection | 4-15% | 12.81% (9.21-16.97) |
| Annual incidence of TB disease after five years since infection | <0.2% | 0.40%* (0.20-0.68) |
| Case fatality in the absence of treatment | 40-70% | 50.60% (26.98-74.27) |
| Mean duration of TB disease in absence of treatment | 1.5-4 years | 4.0 years (2.5-6.8) |
| Reduction in the risk of TB afforded by prior infection | 40-85% | 78.05% (82.60-72.95) |

*The target population represents a vulnerable population with multiple risk factors for TB progression.

**Table S5: Assumptions for TPT regimens**

| **Regimen and parameter input** | **Value** | **Source** |
|---|---|---|
| **9H** | | |
| Number of outpatient visits | 9 | Based on discussion with Ministry of Health staff |
| Number of doses (300mg isoniazid) | 270 | (10) |
| **4R** | | |
| Number of outpatient visits | 4 | Based on discussion with Ministry of Health staff |

| | | |
|---|---|---|
| Number of doses (600mg rifampin) | 120 | (10) |
| **3HP** | | |
| Number of outpatient visits | 3 | Based on discussion with Ministry of Health staff |
| Number of doses (900mg isoniazid/900 mg rifapentine) | 12 | (10) |
| **1HP** | | |
| Number of outpatient visits | 1 | Based on discussion with Ministry of Health staff |
| Number of doses (900mg isoniazid/900 mg rifapentine) | 28 | (11) |
| **Costs (2022 USD)** | | |
| Costs outpatient visit | 3.80 (3.42-5.02) | (12) |
| Costs rifampin dose 300mg | 0.087 | (13) |
| Costs rifapentine dose 150 mg | 0.31 | (13) |
| Costs isoniazid dose 300mg | 0.037 | (13) |

** USD = US dollars.*

### Supplementary exhibits

#### Table S6: (Un)discounted cost per DALY averted for each intervention scenario compared to the base case scenario*

| Strategy | Undiscounted ICER in USD | Discounted ICER in USD |
|---|---|---|
| IGRA/1HP | 298 (73-917) | 708 (293-1892) |
| IGRA/3HP | 242 (45-775) | 596 (225-1,650) |
| IGRA/4R | 248 (57-761) | 607 (246-1,536) |
| IGRA/9H | 300 (82-865) | 702 (296-1,764) |
| TST/1HP | 134 (dominant-512) | 383 (86-1,124) |
| TST/3HP | 64 (dominant-342) | 242 (8-775) |
| TST/4R | 67 (dominant-351) | 250 (10-796) |
| TST/9H | 105 (dominant-439) | 318 (53-949) |

*For optimal decision-making, choice of strategy should be based on the full incremental analysis as presented in the main text. IGRA = Interferon Gamma Release Assays. TST = Tuberculin Skin Test. ICER = Incremental Cost-Effectiveness Ratio. H = isonazid. R = rifampin. P = rifapentine.

#### Table S7: (Un)discounted TB DALY per 1,000 persons associated with each scenario

| Strategy | Undiscounted TB DALY per 1,000 persons | Discounted TB DALY per 1,000 persons |
|---|---|---|
| No TPT | 432 (186-772) | 216 (97-380) |
| IGRA/1HP | 282 (123-521) | 141 (65-253) |
| IGRA/3HP | 287 (125-526) | 143 (65-252) |
| IGRA/4R | 288 (124-523) | 144 (65-257) |
| IGRA/9H | 300 (131-541) | 149 (68-263) |
| TST/1HP | 290 (128-529) | 145 (67-256) |
| TST/3HP | 294 (130-532) | 147 (67-257) |
| TST/4R | 295 (129-541) | 147 (67-261) |
| TST/9H | 306 (134-551) | 153 (70-265) |

* IGRA = Interferon Gamma Release Assays. TST = Tuberculin Skin Test. ICER = Incremental Cost-Effectiveness Ratio. H = isonazid. R = rifampin. P = rifapentine.

#### Table S8: (Un)discounted cost per 1,000 person for each scenario

| Strategy | Undiscounted cost in USD per 1,000 persons | Discounted cost in USD per 1,000 persons |
|---|---|---|
| No TPT | 99,585 (58,552 -157,598) | 70,966 (44,608-105,492) |
| IGRA/1HP | 143,987 (111,634-183,852) | 124,197 (98,889-152,841) |

| | | |
|---|---|---|
| IGRA/3HP | 134,705 (102,462-174,332) | 114,570 (90,930-142,872) |
| IGRA/4R | 135,176 (102,518-178,317) | 114,921 (90,489-143,211) |
| IGRA/9H | 139,155 (106,350-184,177) | 117,965 (93,292-146,538) |
| TST/1HP | 118,631 (83,652-157,452) | 98,410 (72,790-125,969) |
| TST/3HP | 108,375 (76,483-149,531) | 87,824 (64,171-116,243) |
| TST/4R | 108,841 (77,059-152,900) | 88,173 (64,618-116,563) |
| TST/9H | 112,814 (79,274-158,099) | 91,247 (67,210-120,143) |

* IGRA = Interferon Gamma Release Assays. TST = Tuberculin Skin Test. ICER = Incremental Cost-Effectiveness Ratio. H = isonazid. R = rifampin. P = rifapentine.

#### Table S9: Averted DALYs, incremental costs, and ICERs for TST/3HP strategy in comparison to base-case for different ages at testing, duration of incarceration, and time spent since release from prison

| | | **Undiscounted DALYs averted per 1,000 persons** | **Incremental undiscounted costs per 1,000 persons ($)** | **Cost per DALY averted (discounted) ($)** |
|---|---|---|---|---|
| **Age in years** | 25 | 159 ( 62 - 308 ) | 7,350 ( -16,868 - 26,828 ) | 216 (dominant-697) |
| | 30* | 138 ( 55 - 267 ) | 8817 ( -14362 - 27741 ) | 243 (4-760) |
| | 35 | 119 ( 47 - 228 ) | 10,043 ( -11,835 - 28,754 ) | 271 (7-834) |
| | 45 | 84 ( 34 - 159 ) | 13,201 ( -6,482 - 30,699 ) | 367 (42-1,063) |
| | 65 | 29 ( 12 - 52 ) | 22,292 ( 6,699 - 38,399 ) | 1,081 (362-2,596) |
| **Duration of incarceration** | 1 month | 23 ( 8 - 52 ) | 16,885 ( 7,724 - 26,443 ) | 1,716 (610-5,444) |
| | 3 months | 55 ( 21 - 108 ) | 13,546 ( 3,004 - 23,781 ) | 590 (203-1,570) |
| | 6 months | 78 ( 31 - 148 ) | 11,809 ( -536 - 23,862 ) | 392 (105-1,080) |
| | 1 year | 110 ( 43 - 210 ) | 9,904 ( -6,407 - 24,336 ) | 276 (43-806) |
| | 2 years* | 138 ( 55 - 267 ) | 8817 ( -14362 - 27741 ) | 243 (4-760) |
| | 5 years | 155 ( 58 - 319 ) | 8,362 ( -25,054 - 33,582 ) | 271 (dominant-989) |
| | 10 years | 156 ( 56 - 330 ) | 8,213 ( -29,327 - 35,168 ) | 299 (dominant-1,1336) |
| **Time since release** | no time | 142 (57-274) | 7,393 ( -15,441 – 26,026 ) | 208 (dominant-656) |
| | 1 month | 145 ( 57 - 273 ) | 7,367 ( -15,931 – 26,358 ) | 208 (dominant-661) |
| | 3 months* | 146 ( 58 - 276 ) | 8,817 ( -14,362 – 27,741 ) | 243 (4-760) |
| | 6 months | 137 ( 54 - 259 ) | 10,355 ( -12,621 – 29,208 ) | 294 (24-941) |
| | 1 year | 125 ( 48 - 240 ) | 12,104 ( -10,742 – 30,797 ) | 384 (48-1,295) |
| | 2 years | 107 ( 39 - 219 ) | 13,556 ( -9,403 – 31,992 ) | 505 (76-1,902) |
| | 5 years | 91 ( 29 - 192 ) | 15,302 ( -67,84 – 33,672 ) | 638 (115-2,428) |
| | 10 years | 78 ( 24 - 176 ) | 17,315 ( -4,091 – 35,506 ) | 776 (158-2,962) |

*Value used in main analysis. DALY = disability adjusted life years.

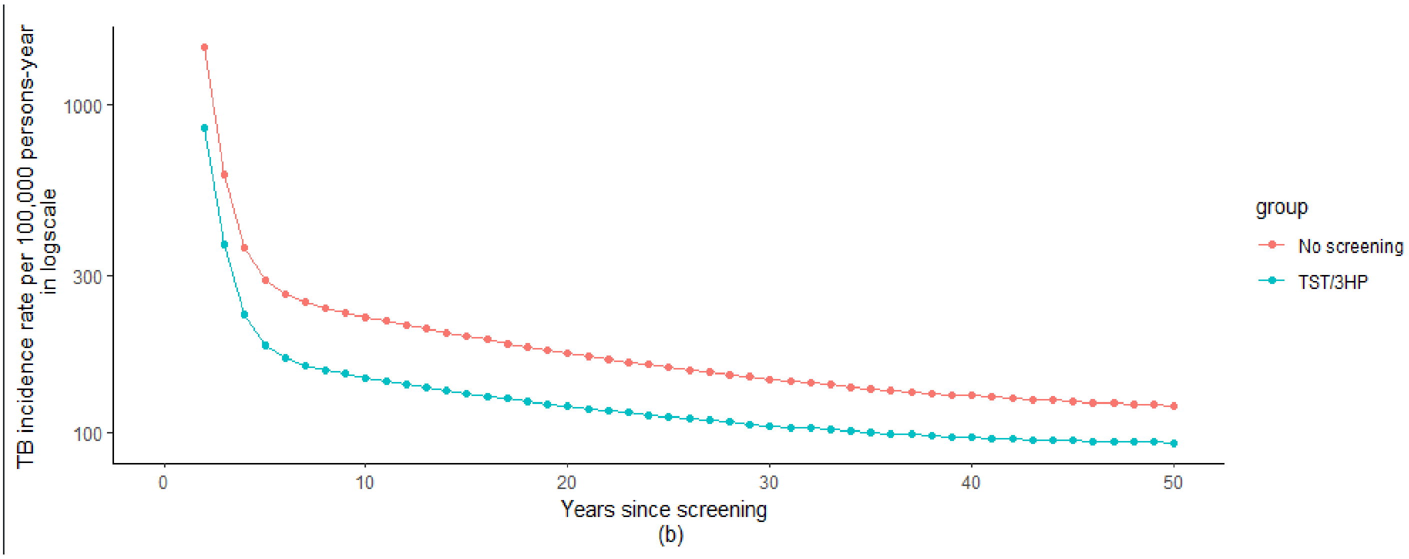


**Figure S1: Annual TB disease incidence rate over 50 years, comparing base case scenario and TST/3HP scenarios.**

### Sensitivity Analysis

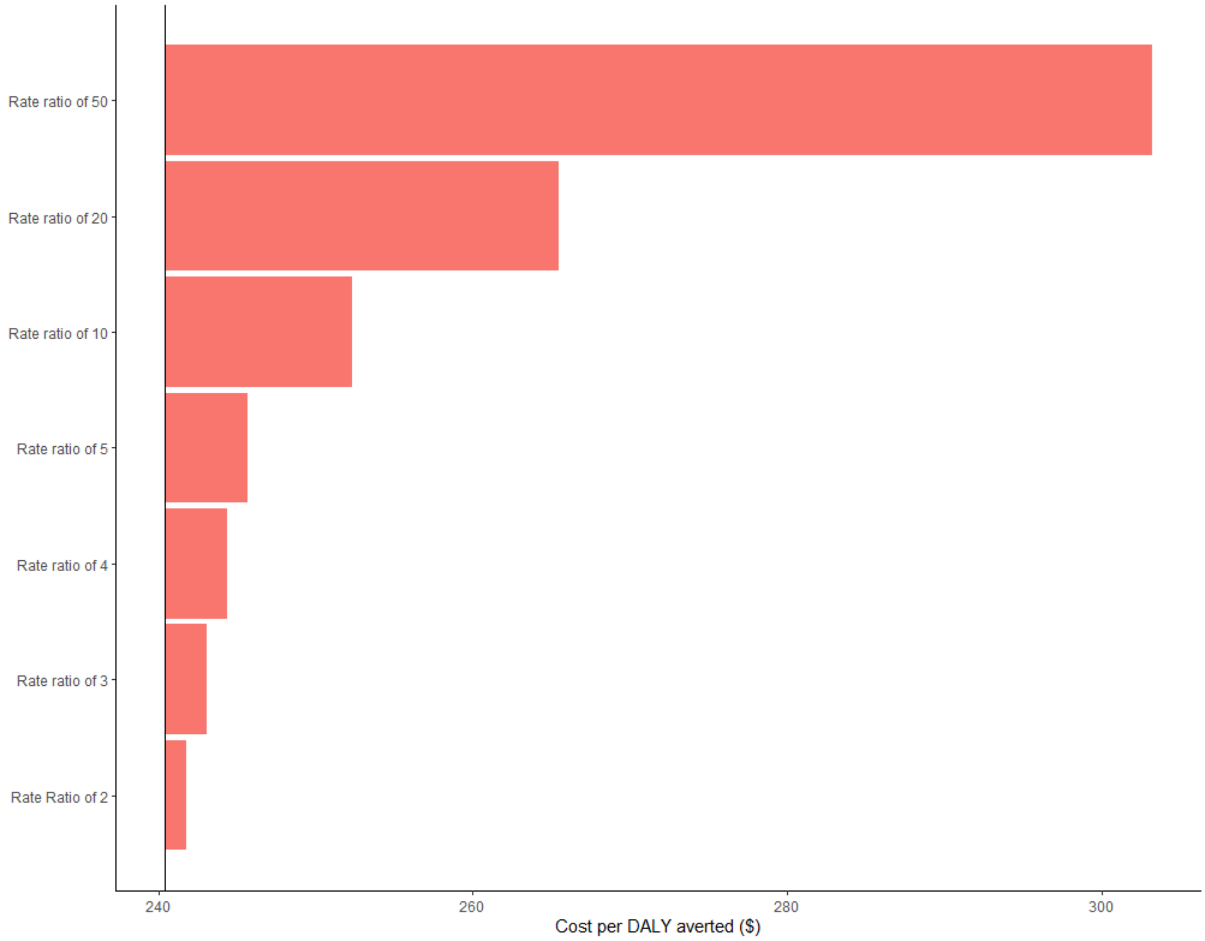


**Figure S2: ICER for TST/3HP vs base case for different values of the rate ratio of the force of infection after prison release. DALY = disability-adjusted life years.**

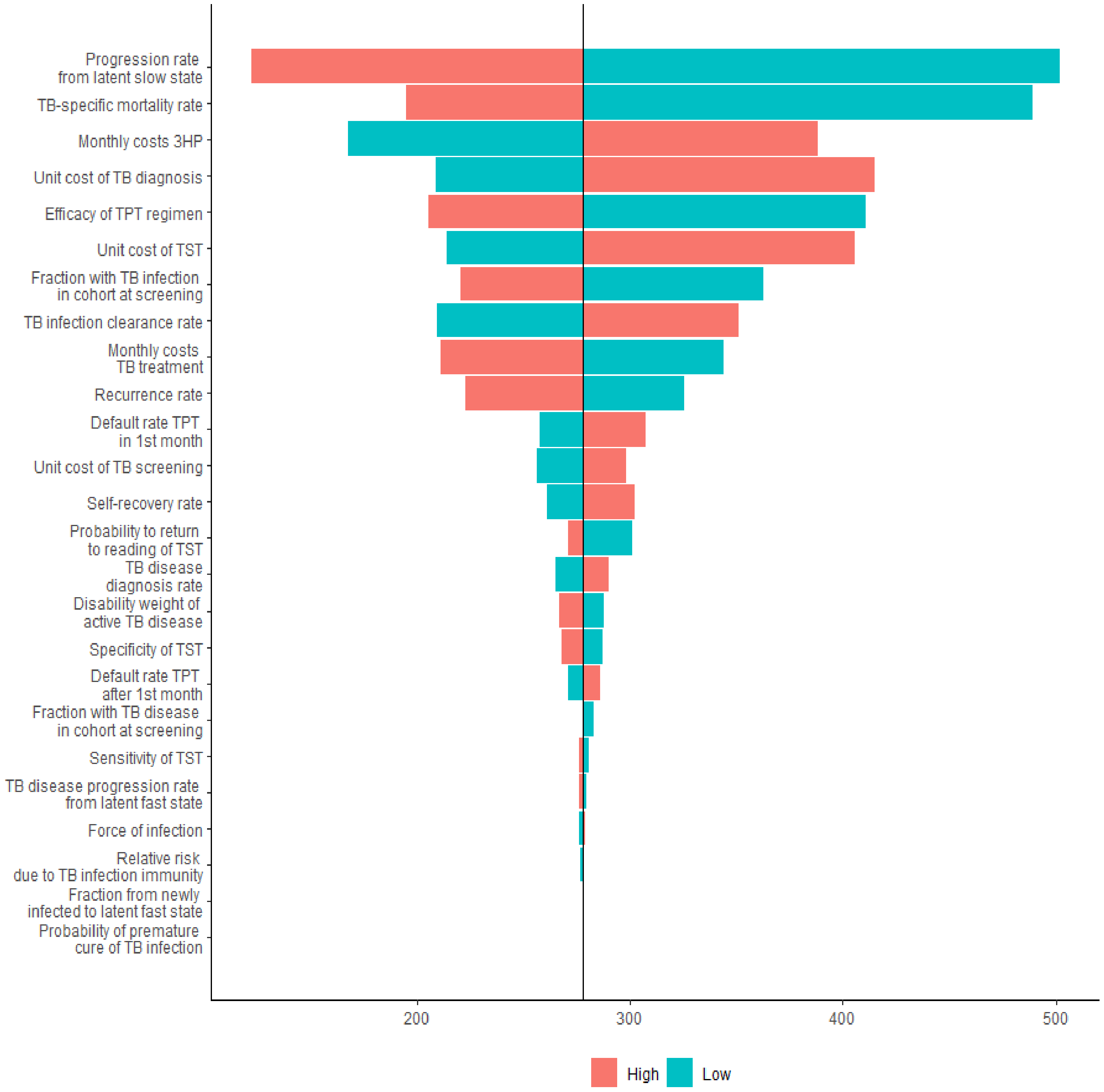


**Figure S3: Tornado diagram showing one-way sensitivity analysis for all parameters used in the model, for the ICER comparing TST/3HP scenario to base-case. TST = Tuberculin Skin Test. H = isoniazid. P = rifapentine. TPT = TB preventive Treatment.**
